# Genotype association of VEGF polymorphism rs2010963 with Strabismus in a Pakistani cohort

**DOI:** 10.1101/2025.10.23.25338643

**Authors:** Muhammad Ahmad Khalid, Noor-ul-ain Amin, Conain Nehal, Muhammad Khaleeq Saqi, Kwang Min Woo, Muhammad Usman Jamil, Maleeha Azam

## Abstract

**Purpose:** This study was conducted to investigate the associations between specific genetic variation in VEGF and the prevalence of strabismus, with particular focus on VEGF polymorphism rs2010963 and whether presence of GG phenotype increases the odds of strabismus compared to GC or CC allele in a Pakistani cohort.

**Patients and methods:** Blood samples from 43 strabismus patients and 170 normal controls (taken from an already published study from our group). Genotyping of the selected single nucleotide polymorphism (SNP) was carried out using polymerase chain reaction restriction fragment length polymorphism (PCR-RFLP) for 43 samples of strabismus. Genotype and allele frequencies were then computed for all patients. Logistic regression was used to evaluate the association between the rs2010963 genotype and the risk of strabismus. A p-value of < 0.05 was considered statistically significant.

**Results:** Of the 43 strabismus patients, 27 (66%) had exotropia, followed by esotropia in 14 patients (34%). Their genotype frequency included 24 (56%) patients with the GG genotype, 14 (33%) with the GC genotype, and 5 (12%) with the CC genotype. Among the 170 normal controls, 50 (29%) had the GG genotype, 60 (35%) had the GC genotype, and 60 (35%) had the CC genotype. Individuals with the GG phenotype had significantly higher odds of having strabismus compared to those with CC genotype (OR = 5.76, 95% CI [2.05–16.20], p = 0.0009). GG phenotype was also associated with higher odds to those with GC phenotype, but the result was not statistically significant (OR = 2.06 [0.96 – 4.39], p = 0.062).

**Conclusion:** This study explored the association of VEGFA polymorphism rs2010963 and strabismus in a Pakistani cohort. There is a higher prevalence of the GG genotype amongst patients with strabismus compared to normal controls, and a significant association between rs2010963 and strabismus.

## Introduction

Strabismus, defined as a misalignment of the eyes, is among the most prevalent pediatric ocular disorders, affecting approximately 2–4% of the global population, with similar or higher rates reported in Pakistani children.^1,2^ This condition can lead to amblyopia, reduced stereopsis, and, in severe cases, permanent vision loss.^3,4^ Beyond visual consequences, strabismus has been shown to negatively affect psychosocial well-being, self-esteem, and quality of life.^2,5,6^ The pathophysiology of strabismus is complex, encompassing disruptions in extraocular muscle function, anomalies in orbital connective tissues, and neural control deficits.^3,7^ While environmental factors such as premature birth, low birth weight, and perinatal insults contribute to risk, a substantial heritable component is well documented.^4,8^

Genetic studies have highlighted the polygenic and heterogeneous nature of strabismus. Twin and family studies suggest a heritability of up to 70% in monozygotic twins and 30–40% in dizygotic twins, with several candidate loci, such as the STBMS1 locus, implicated in familial cases.^3,5^ Advances in exome sequencing and linkage analyses have identified a spectrum of genes related to ocular muscle development, cranial nerve innervation, and cortical visual processing pathways.^4,8^ However, the molecular mechanisms linking these genetic variants to strabismus remain incompletely understood.

Among the genes of interest, vascular endothelial growth factor A (VEGFA) has attracted attention for its role in angiogenesis, myogenesis, and neuroprotection.^9^ VEGFA is a key cytokine regulating vascular permeability, endothelial cell proliferation, and extracellular matrix remodeling, all processes relevant to the development and maintenance of extraocular muscles.^10^ Importantly, differential expression of VEGFA and its receptors has been observed in strabismic extraocular muscles, suggesting a role in altered muscle or nerve remodeling.^11,12^

Single-nucleotide polymorphisms (SNP) in the VEGFA gene, including rs2010963 (commonly referred to as +405G>C or −634G>C), have been shown to influence VEGFA expression levels in retinal and other vascular tissues.^10^ The rs2010963 polymorphism has been linked to several angiogenesis-related diseases such as diabetic retinopathy, retinopathy of prematurity, and coronary artery disease, suggesting its functional relevance.^9,13^ Given VEGFA’s importance in extraocular muscle physiology and neurovascular interactions, VEGFA polymorphisms may represent plausible genetic contributors to strabismus risk.

To date, little is known about the association between VEGFA genetic variants and strabismus in South Asian populations, including Pakistan, where the genetic and environmental profiles may differ from previously studied cohorts. Therefore, this study aimed to investigate the association between the VEGFA rs2010963 polymorphism and strabismus in a Pakistani cohort. Understanding whether specific VEGFA variants, such as rs2010963, are associated with an increased risk of strabismus could help in better understanding its genetic etiology.

## Material and methods

In this case-control study, a total of 43 patients with strabismus were recruited, and blood samples were collected for analysis. For the control group, genotype data from 170 healthy individuals previously published by our group were used as population controls.^11^

The genotypes of the selected SNP, rs2010963, were determined using PCR-RFLP, where the amplified products were digested with the BsmF1 restriction enzyme. This enzyme cuts the DNA in a way that allows a clear distinction between the GG, GC, and CC genotypes based on the resulting fragment sizes observed on agarose gel **(Figure 1)**.

**Figure 1.**
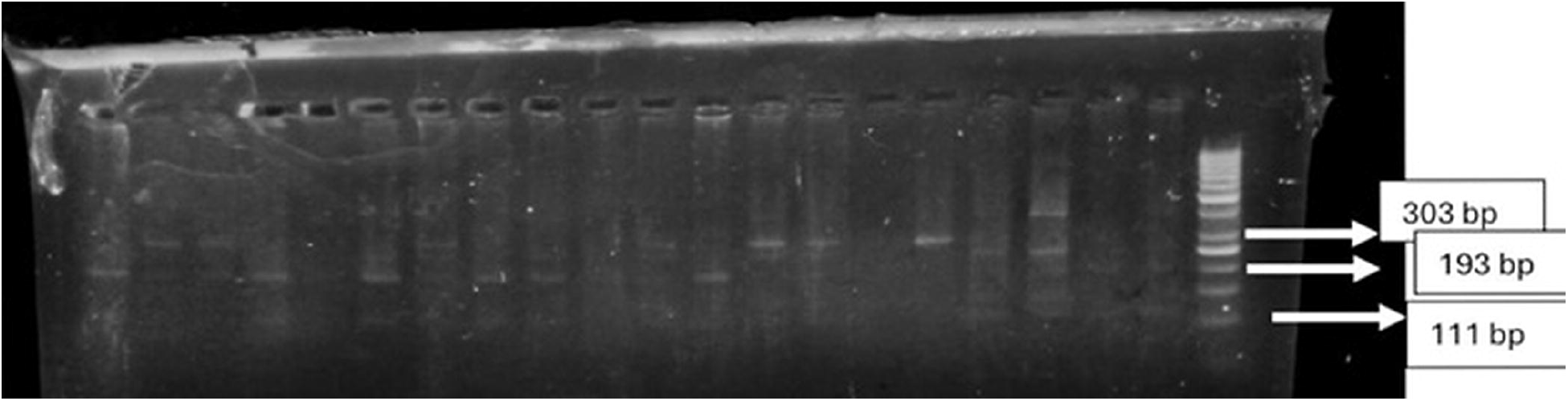
The gel shows the results of RFLP digestion. The presence of 111 bp and 193 bp bands indicates the GG genotype, resulting from successful restriction enzyme digestion. In contrast, the presence of an uncut 303 bp band corresponds to the CC genotype, indicating no restriction site for the enzyme. When all three are present, this represents a GC genotype (such as the one marked on the right of the gel)

The study adhered to the principles of the Declaration of Helsinki and was approved by the Ethics Review Board of the Department of Biosciences, COMSATS University Islamabad (CUI-Reg/Notif-452/20/526). Written informed consent was obtained from all participants.

### Statistical Analysis

Genotype and allele frequencies were calculated for cases and controls, and associations between VEGFA rs2010963 genotypes and strabismus were evaluated using logistic regression, with odds ratios (OR) and 95% confidence intervals (CI). A p-value of <0.05 was considered statistically significant. The analyses were performed in Python (version 3.11).

## Results

### Genotype Distribution

A total of 43 patients with strabismus and 170 healthy controls were included in the analysis. **Table 1** includes the demographic data of the patients and controls sampled. Out of the 43 strabismus patients, 41 had information on the specific type of strabismus they had. Among these, Exotropia was the most common type, seen in 27 patients (66%), followed by Esotropia in 14 patients (34%). The genotype distribution of VEGFA rs2010963 in strabismus patients was as follows: 24 individuals (56%) had the GG genotype, 14 individuals (33%) had the GC genotype, and 5 individuals (12%) had the CC genotype. In contrast, among the 170 controls, 50 individuals (29%) were GG, 60 (35%) were GC, and 60 (35%) were CC genotype carriers.

**Table 1.** Demographic data and descriptive features. Table 1 shows demographics in the strabismus subjects and controls. (SD= standard deviation)

|  | Strabismus Subjects (n=43) | Controls (n=170) |
| --- | --- | --- |
| Sex Male (%) | 22 (51.2%) | 79 (46.5%) |
| Age in years (SD) | 18.42 (8.40) | 40.55 (9.14) |

### Genotype-Based Association

Logistic regression analysis revealed that individuals with the GG genotype had significantly higher odds of developing strabismus compared to those with the CC genotype (OR□=□5.76, 95% CI□2.05–16.20, p□=□0.0009). When comparing the GG genotype to the GC genotype, the odds ratio was 2.06 (95% CI 0.96–4.39), though this association did not reach statistical significance (p□=□0.062). No statistically significant difference was observed between the GC and CC genotypes (OR□=□1.23, 95% CI 0.44–3.44, p□=□0.68) **(Table 2)**.

**Table 2.** Frequency distribution and logistic regression analysis of VEGFA rs2010963 genotypes and alleles in strabismus cases and controls. Table 2 presents the frequency distribution and logistic regression-based association analysis of the VEGFA rs2010963 polymorphism in 43 strabismus cases and 170 healthy controls. Genotype counts and frequencies are shown for each group, along with odds ratios (ORs) and 95% confidence intervals (CIs) calculated using logistic regression. Comparisons were performed using the CC genotype as the reference group for genotype models, and the C allele as the reference for allele models. The allele-based association is derived by counting the total G and C alleles among cases and controls. Percentages reflect the proportion of each genotype or allele within its respective study group.

| Comparison | Strabismus Cases (n=43) | Controls (n=170) | Frequency in Cases (%) | Frequency in Controls (%) | OR (95% CI) | p-value |
| --- | --- | --- | --- | --- | --- | --- |
| **GG vs GC** | 24 (GG), 14 (GC) | 50 (GG), 60 (GC) | 55.8% (GG), 32.6% (GC) | 29.4% (GG), 35.3% (GC) | 2.06 (0.96–4.39) | 0.062 |
| **GG vs CC** | 24 (GG), 5 (CC) | 50 (GG), 60 (CC) | 55.8% (GG), 11.6% (CC) | 29.4% (GG), 35.3% (CC) | 5.76 (2.05–16.20) | 0.0009 |
| **GC vs CC** | 14 (GC), 5 (CC) | 60 (GC), 60 (CC) | 32.6% (GC), 11.6% (CC) | 35.3% (GC), 35.3% (CC) | 1.23 (0.44–3.44) | 0.68 |
| **Allele G vs C** | 62 G, 24 C (86 total alleles) | 160 G, 180 C (340 total alleles) | 72.1% G | 47.1% G | 2.91 (1.73–4.87) | 5.55×10<sup>-4</sup> |

### Allele-Based Association

An allelic association analysis was performed to further evaluate risk, considering 86 alleles from cases and 340 alleles from controls. The frequency of the G allele was 72.1% among strabismus patients compared to 47.1% among controls. Pearson’s chi-square test (1 df) yielded a value of χ^2^□=□16.25, corresponding to an allelic OR of 2.91 (95% CI□1.73–4.87) (p□=□5.55□×□10□□) **(Table 2)**. This significant enrichment of the G allele in strabismus cases supports the genotype-based finding of an increased risk associated with the rs2010963-G variant.

## Discussion

In this small pilot study, we investigated the association between the VEGFA rs2010963 polymorphism and strabismus in a Pakistani cohort. Our findings demonstrate that the GG genotype of rs2010963 has significantly higher odds of contributing to strabismus compared to healthy controls, leading to more than five-fold increased risk of strabismus relative to the CC genotype. Furthermore, allelic analysis revealed a significantly higher frequency of the G allele among cases, consistent with its potential role as a risk allele.

These observations are noteworthy given the established role of VEGFA in angiogenesis, myogenesis, and neurovascular development.^9^ VEGFA has been implicated in the physiology of extraocular muscles and their vascular supply.^10^ The altered VEGFA expression could plausibly affect muscle contractility or remodeling, predisposing individuals to strabismus.^9,10^ Prior studies have reported downregulation of VEGFA signaling components in strabismic extraocular muscles, suggesting a pathogenic link between vascular growth factor pathways and ocular alignment disorders.^1,14,15^

The rs2010963 polymorphism has previously been studied in relation to other vascular-related ocular conditions, including diabetic retinopathy, retinopathy of prematurity, and age-related macular degeneration.^9,11,13^ Experimental studies have shown that this SNP influences VEGFA gene expression levels in various tissues, including the retina, where the C allele is generally associated with higher expression.^10^ In contrast, the lack of VEGFA expression associated with the G allele could lead to reduced VEGFA signaling.^10^ As the VEGFA is necessary for the neurogenic development of the extraocular muscles, this could lead to increased susceptibility to strabismus, as observed in our study compared to controls.^12^

Overall, strabismus has a complex genetic background, with several genes described in the literature reflecting the functional heterogeneity and multifactorial nature of the disorder.^3,8^ However, confirming these findings across studies has been challenging due to differences in clinical presentation and genetic architecture among populations.^3,8^ Our results suggest that VEGFA could be an important genetic factor in strabismus, especially in South Asian populations, where unique genetic and environmental backgrounds may influence disease risk. Since VEGFA polymorphism frequencies can vary among different ethnic groups, larger studies involving more diverse populations are needed to validate and confirm these observations.^16^

Limitations of our study include the use of controls from a prior study and a relatively small sample size, which limits the power to detect associations with small effect sizes and may lead to inflated odds ratios. Second, our study was limited to a single SNP for analysis, and other VEGFA variants or haplotypes, in combination with environmental or epigenetic factors, could also influence the strabismus phenotype. Larger and more diverse studies, along with lab research on VEGFA in eye muscles, are needed to better understand these results and guide future treatments.

## Conclusion

Our study is the first to report a significant association between VEGFA rs2010963 and strabismus in a Pakistani population, linking the GG genotype with a higher risk for strabismus. Both genotype and allele results suggest that VEGFA may play a role in genetic risk for this condition, thereby supporting the involvement of angiogenic pathways in strabismus.

## Data Availability

All data produced in the present work are contained in the manuscript

## Disclosure

The author(s) report no conflicts of interest in this work.

## Notes

### Competing Interest Statement

The authors have declared no competing interest.

### Funding Statement

This study did not receive any funding

### Author Declarations

Ethics Review Board of the Department of Biosciences, Commission on Science and Technology for Sustainable Development in the South University Islamabad granted the approval of the work.

